# Early behavior of Madrid Covid-19 disease outbreak: A mathematical model

**DOI:** 10.1101/2020.03.30.20047019

**Authors:** Daniel García-Iglesias, Francisco Javier de Cos Juez

## Abstract

**Introduction:** Madrid Covid-19 disease outbreak started on 28 February 2020 and since then it became the main Covid-19 disease cluster in Spain. On 26 March 2020, a total of 17166 cases were already reported, with 2090 deaths. Globally a R_0_ index of 2-3 has been reported. We aimed to build an experimental mathematical model that help to analyze the early characteristics of Madrid Covid-19 disese outbreak and to explore the actual R0 index on Madrid Covid-19 outbreak.

**Material and Methods:** A simulated mathematical model was built, based on a SIR epidemiological model and the reported characteristics of Wuhan Covid-19 disease outbreak. Monte Carlo simulations were further done to estimate the R_0_ value over time in the Madrid Covid-19 disease outbreak.

**Results:** Mean estimated R0 value along the early period is of 2.22 (+/-1.21 SD). A significant increase of 0.093 (+/-0.037, p=0.025) in R0 value each day of outbreak is found.

**Conclussions:** Our proposed Mathematical Simulation model may be useful to evaluate early characteristics of this outbreak. The present work is the first reported estimation of R0 value in the Spanish Madrid Covid-19 outbreak, with similar results to the previous reported in the Wuhan outbreak, although it may suggest a slightly increase on R0 along time.

## Introduction

### Epidemic Background

On 31 December 2019, a comunity-adquired pneumonia of unknown ethiology was first reported in Wuhan city. On 2 January 2020 it was first published its relation with a Coronaviridae virus, called SARS-CoV-2(1). During the first two months of epidemic, a total of 79384 cases were reported in China, with 2838 deaths (Lethal Index of 3.57%(2). On 13 January 2020 the first case outside China was reported in Thailand, corresponding to an imported case from Wuhan and in the following days, imported cases from this region were also reported in Japan and Republic of Korea(3).

During the first 2 months of epidemia, a total of 85403 cases were globally reported, with a total of 2924 deaths and a Letality index of 3.42%(2).

In Spain, the first reported cases occurred on 31 January and 9 February 2020. They were two cases adquired on Germany and United Kingdom respectively(4). And the first important clusters with local transmision were reported on 28 February 2020 in the Canary Islands, Andalucia, Valencia and Madrid(5). From them, the main cluster was Madrid, where on 26 March 2020, a total of 17166 cases were already reported, with 2090 Covid-19 deaths.

On 13 March 2020, the day before Public Health Measures (PHM) were taken, there were 4209 cases reported in Spain (47.28% of them −1990 cases-from Madrid). This PHM were based on mobility restrictions. Covid-19 disease: Epidemiological Characteristics

Covid-19 disease is caused by a Coronaviridae virus called SARS-CoV-2[Chan2019]. It is transmitted from direct contact with respiratory drops, hands and contaminated fomites. Mean incubation period is of 5 days and time to peak symptoms is of 3 days(6,7). Time to recovery from symptoms is of 14 days, although it can take up to 30 days (6,7).

According to the early Wuhan outbreak data, the Reproduction Number (R_0_) is reported to be between 2 and 3 [Li, Wu, Riou]. Although it may change over time and it can be reduced with PHM based on mobility restrictions, as it has been seen in Wuhan(8,9).

### Objectives

To describe the early characteristics of Covid-19 outbreak in Madrid.

To build a Mathematical model that fits the early findings of Covid-19 infection on Madrid’s outbreak.

## Material and Methods

### Study data-set

Data used for this paper was obtained directly from Spanish Government reported cases. This data is available online at the Spanish Health Ministry webpage (10). Analyzed data corresponds to the first 15 days of Madrid Covid-19 disease outbreak (26 February to 11 March 2020).

### Mathematical model

Based on the Spanish Government reported cases, we built a stochastic transmission model, based on a classical SIR epidemiological model (11), as can be seen on figure 1. Basically, it divides individuals into four different subgroups: Susceptible, Exposed, Infected and Removed (recovered, isolated or death).

**Figure 1.**
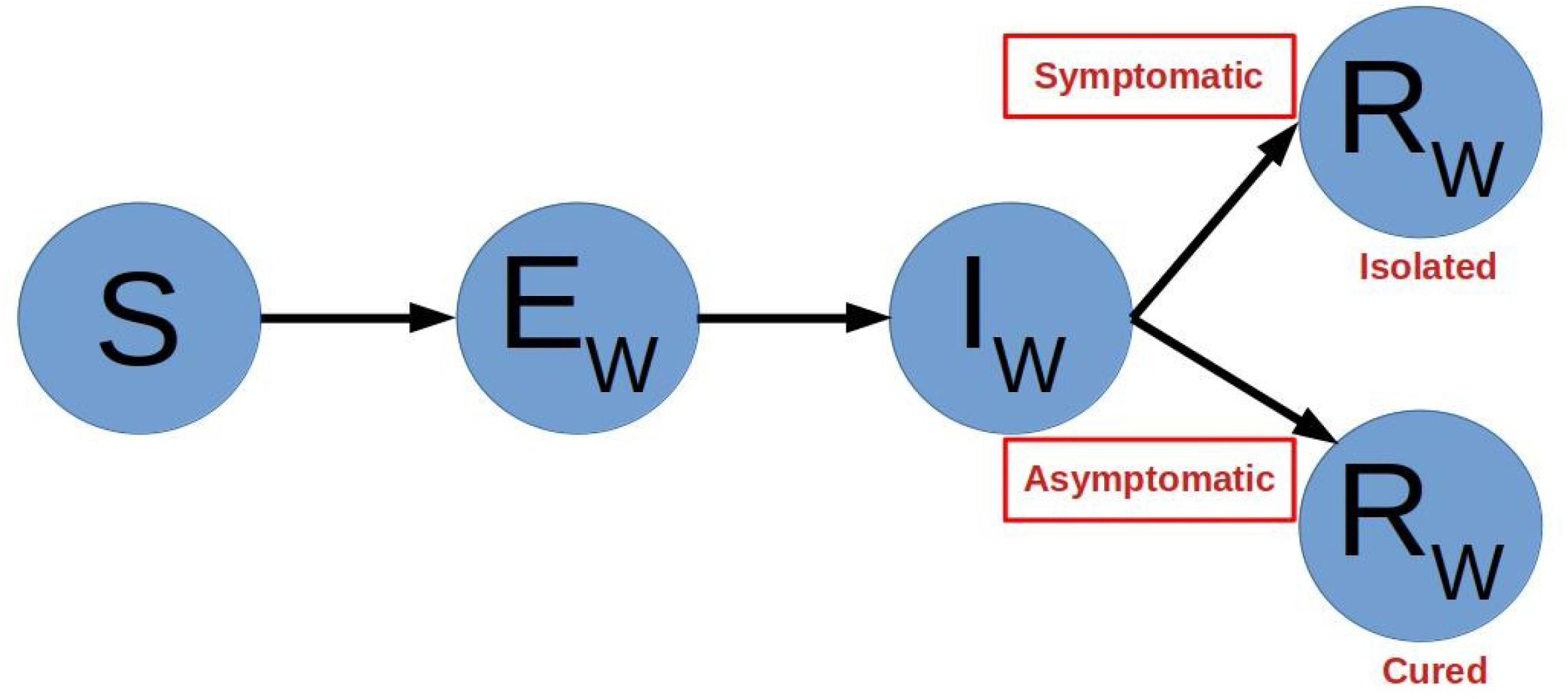
Proposed SIR model for the simulations.

The incubation period was assumed to be Normally distributed, with a mean of 5 days and a standard deviation (SD) of 2 days. The time to the start of symptoms and the peak of the disease is normally distributed, with a mean of 3 days and a SD of 1 day. Once a patient is infected (turns into an Infected individual), the probability to transmit the infection on a certain day (*R*_*n*_) is based on the estimated *R*_0_, with a peak probability on the peak day (*p*) of symptoms:

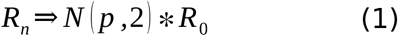

The probability for developing symptoms is fixed on 0.7, and in those symptomatic patients, the time to diagnosis is Normally distributed, with a mean of 8 days and a SD of 2 days. Once a patient is diagnosed, it is supposed to be removed from the general population (turns into a Removed individual) and thus its probability to infect other individual is stopped. If a patient is asymptomatic and thus no diagnosed, it still can infect other individuals up to 12 days (with a SD of 2 days) after the supposed peak of symptoms. All of this parameters introduced in the model are shown in table 1.

**Table 1.** Description of variables and values included in the Mathematical model.

| Variable | Description | Values |
| --- | --- | --- |
| $i$ | Day of infection | |
| $T_i$ | Time from infection to beginning of symptoms (incubation) | 5 (± 2) days |
| $p$ | Peak day of symptoms | |
| $T_p$ | Time from beginning of symptoms to $p$ day (peak of symptoms) | 3 (± 1) day |
| $n_s$ | Days with possible risk for infection transimission (in a symptomatic patient) | 5 (± 2) days |
| $n_a$ | Days with possible risk for infection transimission (in an asymptomatic patient) | 15 (± 2) days |
| $R_n$ | Infection risk in each time epoch, as described in formula 1 | Formula 1 |

The Mathematical model was built with R (12) and it can be downloaded online from our repository [cite].

### Simulations and Statistic analysis

With sequential Monte Carlo simulations, based on different R_0_ values over time, the number of infected and diagnosed individuals at the beginning of the Madrid Covid-19 outbreak are estimated for each time epoch. This estimated diagnosed individuals are compared with the reported cases of Covid-19 disease and thus, the probability of each R_o_ value is calculated in each time epoch. R_0_ values are expressed as most probably, 95% confidence interval and 90 percent confidence interval.

To asses if the estimated R_0_ is stable along time, initial estimated R_0_ values are compared with the final values (first to 3^rd^ day against 13^th^ to 15^th^ day of outbreak). And finally all estimated R_0_ values for each time epoch are regressed against each day of outbreak.

## Results

Mean estimated R_0_ value along the early period is of 2.22 (+/-1.21 SD). A temporal representation of the obtained R_0_ value is shown in figure 2, and temporal results with the confidence interval is also shown in table 2.

**Table 2.** Evolution of R_o_ value along the first days of Covid-19 Madrid outbreak.

|  | 26 February | 27 February | 28 February | 29 February | 1 March | 2 March | 3 March | 4 March |
| --- | --- | --- | --- | --- | --- | --- | --- | --- |
| $R_0$ | 2.3 | 1.9 | 0.7 | 1.6 | 2.7 | 2.4 | 3.4 | 2.6 |
| 95% CI | 1.9 – 3.6 | 1.4 – 2.3 | 0.4 – 0.8 | 1.3 – 2 | 2.4 – 2.8 | 2.2 – 2.6 | 2.9 – 3.8 | 2.3 – 2.8 |
| 90% CI | 1.5 – 4.6 | 1.1 – 2.8 | 0.4 – 1 | 1.2 – 2.4 | 2.3 – 3.3 | 1.9 – 2.9 | 2.9 – 4 | 2 – 2.8 |

**Table 2.** Evolution of $R_0$ value along the first days of Covid-19 Madrid outbreak.
|  | 5 March | 6 March | 7 March | 8 March | 9 March | 10 March | 11 March |
| --- | --- | --- | --- | --- | --- | --- | --- |
| $R_0$ | 1.6 | 2.5 | 2.2 | 2.4 | 3 | 3.2 | 3.3 |
| 95% CI | 1.6 – 1.6 | 2.4 – 2.6 | 2.2 – 2.3 | 2.4 – 2.4 | 3 – 3.1 | 3.1 – 3.3 | 3.3 – 3.4 |
| 90% CI | 1.4 – 1.8 | 2.3 – 2.8 | 2 – 2.4 | 2.3 – 2.5 | 2.9 – 3.1 | 3 – 3.4 | 3.2 – 3.5 |

**Figure 2.**
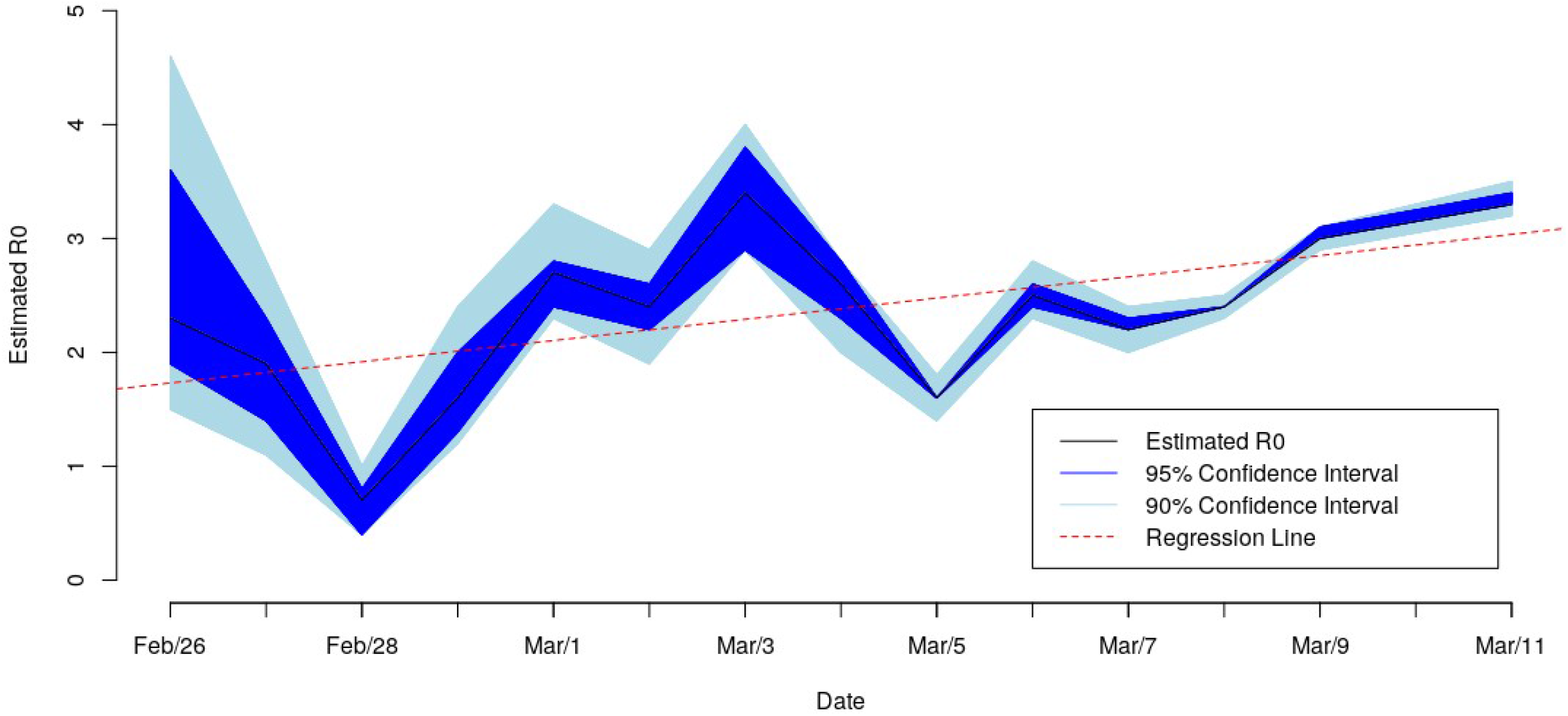
Estimated R_0_ values for each time epoch along the first 15 days of Madrid Covid-19 disease outbreak. Values are expressed as most likely (black line), 95% confidence interval (dark blue) and 90% confidence interval (light blue). The regression line for the estimated R_0_ value against day of outbreak (orange dashed line).

During this analyzed period of time the regression analysis shows a significant increase of the R_0_ value along days. An increase of 0.093 (+/-0.037, p=0.025) in R_0_ value each day of outbreak is found.

## Discussion

Simulated mathematical models are useful to asses the epidemic characteristics and evaluate possible effects of different PHM interventions. Moreover, they are of special interest in early stages of an outbreak, when no enough information is still available to make decisions. In this sense, the proposed model can be used and adapted to evaluate the evolution of Covid-19 disease outbreak and to monitor the results of PHM undertaken by the governments.

As described in the present work, the R_0_ value in Madrid’s Covid-19 disease outbreak (Spanish main Covid-19 disease outbreak) is similar to the reported in previous documents, corresponding to Wuhan outbreak (6,7). This R_0_ value is about 2-3 at the beginning, although our data suggest it can increase along the outbreak if no PHM are taken.

In previous reports, PHM based on mobility restrictions can reduce this R_0_ value, leading to a disease control and finally to a possibility to overcome this situation. In this sense further investigation will be necessary to evaluate weather or not, this mobility restrictions can lead to a reduction in R_0_ value. This effect in outbreak reduction would be of special importance to plan future decisions and to monitor their effect.

## Conclusions

Our proposed Mathematical Simulation model may be useful to evaluate early characteristics of this outbreak and the results of further PHM.

The present work is the first reported estimation of R_0_ value in the Spanish Madrid Covid-19 outbreak. It is similar to the previous reported in the Wuhan outbreak, although it may suggest a slightly increase on R_0_ along time.

Whether or not PHM based on mobility restrictions will be able to reduce R_0_ value in Covid-19 Madrid outbreak is still not known, and thus, further investigation in this sense will be needed.

## Data Availability

All data is available in the Spanish Health minister webpage.

https://www.mscbs.gob.es/profesionales/saludPublica/ccayes/alertasActual/nCov-China/situacionActual.htm

## References

1. Chan JF-W, Kok K-H, Zhu Z, Chu H, To KK-W, Yuan S, et al. Genomic characterization of the 2019 novel human-pathogenic coronavirus isolated from a patient with atypical pneumonia after visiting Wuhan. Emerg Microbes Infect. 2020;9(1):221–36.

2. World Health Organization. Novel Coronavirus(2019-nCoV). Situation report 40. 29 February 2020. [Internet]. 2020 [cited 2020 Mar 20]. Available from: https://www.who.int/docs/default-source/coronaviruse/situation-reports/20200229-sitrep-40-covid-19.pdf?sfvrsn=849d0665_2

3. World Health Organization. Novel Coronavirus(2019-nCoV). Situation report 1. 21 January 2020. [Internet]. 2020 [cited 2020 Mar 20]. Available from: https://www.who.int/docs/default-source/coronaviruse/situation-reports/20200121-sitrep-1-2019-ncov.pdf?sfvrsn=20a99c10_4

4. Centro de Coordinación de Alertas y Emergencias Sanitarias (Ministerio de Sanidad de España). Actualización número 21. Neumonía por nuevo coronavirus COVID-19. [Internet]. 2020 [cited 2020 Mar 20]. Available from: https://www.mscbs.gob.es/profesionales/saludPublica/ccayes/alertasActual/nCov-China/documentos/Actualizacion_21_COVID-19_China.pdf

5. Centro de Coordinación de Alertas y Emergencias Sanitarias (Ministerio de Sanidad de España). Actualización número 36. Neumonía por nuevo coronavirus COVID-19. [Internet]. 2020 [cited 2020 Mar 20]. Available from: https://www.mscbs.gob.es/profesionales/saludPublica/ccayes/alertasActual/nCov-China/documentos/Actualizacion_36_COVID-19.pdf

6. Guan W, Ni Z, Hu Y, Liang W, Ou C, He J, et al. Clinical Characteristics of Coronavirus Disease 2019 in China. N Engl J Med [Internet]. 2020 Feb 28 [cited 2020 Mar 30]; Available from: https://www.nejm.org/doi/10.1056/NEJMoa2002032

7. World Health Organization. Report of the WHO-China Joint Mission on Coronavirus Disease 2019 (COVID-19). [Internet]. 2020 [cited 2020 Mar 20]. Available from: https://www.who.int/docs/default-source/coronaviruse/who-china-joint-mission-on-covid-19-final-report.pdf

8. Huang LL, Shen SP, Yu P, Wei YY. Dynamic basic reproduction number based evaluation for current prevention and control of COVID-19 outbreak in China. Zhonghua Liu Xing Bing Xue Za Zhi Zhonghua Liuxingbingxue Zazhi. 2020 Mar 1;41(4):466–9.

9. Zhang S, Diao M, Yu W, Pei L, Lin Z, Chen D. Estimation of the reproductive number of novel coronavirus (COVID-19) and the probable outbreak size on the Diamond Princess cruise ship: A data-driven analysis. Int J Infect Dis IJID Off Publ Int Soc Infect Dis. 2020 Feb 22;93:201–4.

10. Ministerio de Sanidad. Enfermedad por nuevo coronavirus, COVID-19. Situación actual. [Internet]. [cited 2020 Apr 20]. Available from: https://www.mscbs.gob.es/profesionales/saludPublica/ccayes/alertasActual/nCov-China/situacionActual.htm

11. Kermack WO, McKendrick AG, Walker GT. A contribution to the mathematical theory of epidemics. Proc R Soc Lond Ser Contain Pap Math Phys Character. 1927 Aug 1;115(772):700–21.

12. Team RDC. R: A language and environment for statistical computing [Internet]. Vienna, Austria: R Foundation for Statistical Computing; 2008. Available from: http://www.r-project.org

